# Investigating longitudinal changes in lumbar discs in a two-year interval using quantitative MRI

**DOI:** 10.1101/2025.05.05.25326903

**Authors:** L. Tugan Muftuler, Brendan P. Bych, Jordan A. Gliedt, Jeffrey A. King, John P. Wanner, Megan M. Bellman, Pranjal Srivastava

## Abstract

**Study design:** Longitudinal study using quantitative MRI techniques.

**Purpose:** Disc degeneration leads to a cascade of pathological changes in the spine that might initiate pain. It is particularly important to monitor patients who already have chronic low back pain (CLBP) to observe accelerated degeneration in adjacent segments. Therefore, our goal was to monitor the progression of disc degeneration in CLBP patients over a two-year interval using quantitative MRI techniques.

**Overview of Literature:** Earlier studies investigated age-related changes in intervertebral discs using cross sectional designs or follow-ups with long time intervals. Furthermore, they used qualitative assessment of MRIs.

**Methods:** Oswestry disability index (ODI) questionnaire was administered, and MRI data were acquired from 12 patients with chronic low back pain. Median follow-up period was 2 years. Disc height loss was measured and changes in disc physiology and structure were probed using *T1ρ* relaxation and diffusion MRI. Longitudinal changes in MRI metrics, ODI and their correlations were analyzed.

**Results:** All lumbar discs had significant disc height loss between the baseline and follow-up scans. However, changes in two discs were not significant if stringent Bonferroni correction was used for multiple comparisons. Also, *T1ρ* or diffusion values did not change significantly at the group level during this period. There were strong positive correlations between DHI and ADC changes in L4/L5, between ADC and *T1ρ* changes in L2/L3, and between DHI and *T1ρ* changes in L5/S1. More importantly, these quantitative metrics enabled us to identify changes in disc structure and physiology in individual patients.

**Conclusion:** Quantitative MRI provided complementary information to assess longitudinal changes in IVDs. Although changes were not clearly seen by visual assessments, some participants had significant disc height loss. The quantitative approaches presented here might aid clinicians with more accurate evaluation of the stage of IVD degeneration and monitor its progression.

## INTRODUCTION

Disc degeneration is a part of normal aging processes and common in asymptomatic individuals [1, 2]. However, evidence indicates a strong association between chronic low back pain (CLBP) and a cascade of pathological changes in the spine initiated by disc degeneration [2]. Previous studies have shown that the progression and sequelae of degeneration vary across individuals, and the factors influencing progression are not fully understood [1]

The ability to monitor the progression of disc degeneration could aid in, for instance, detecting adjacent segment degeneration, a frequent complication after spinal arthrodesis [3, 4]. It may also help monitor CLBP patients for the progression of disc degeneration at different levels. This could prompt the physician to take proactive measures before more severe symptoms develop in new segments.

Earlier studies investigating longitudinal changes in intervertebral discs used follow-up intervals ranging from 4 to 17 years [5–10]. These studies typically employed qualitative measures, such as the Pfirrmann classification [11], disc herniation, Modic changes [12], and disc space narrowing. While these studies provided valuable information, cross-sectional designs or long time intervals in longitudinal designs limit our understanding of faster degenerative processes. Moreover, qualitative assessments might be insufficient to discern accelerated degenerative processes from normal age-related changes over shorter periods.

Several groups explored the use of quantitative MRI metrics to augment clinical evaluation [13–19]. A study published in 2015 reported a critical threshold of disc height loss, after which quantitative MRI values indicate an accelerated decline [13, 15]. A disc crossing that threshold might lead to clinically important conditions such as spinal stenosis, intervertebral joint instability, radiculopathy, or myelopathy [1, 20, 21].

Therefore, the objective of this study was to assess degenerative changes in the adult lumbar spine using quantitative MRI metrics in a two-year interval. We report both typical trajectories of degeneration during this period and individual variations. This exploratory study provides further evidence that visual inspection of MRIs may not be adequate for precise assessment of the progression of disc degeneration, and a set of quantitative MRI metrics could help capture complex physiological changes in degenerating discs more accurately.

## MATERIALS AND METHODS

### Ethics Statement

This study is conducted in compliance with the principles of the Declaration of Helsinki. The study was approved by the Institutional Review Board (PRO15564) and written consents were obtained from the participants.

### Study Design

This work is part of a larger study conducted between 2012 and 2016, which investigated degenerative changes in the lumbar spine and their associations with chronic low back pain (CLBP). Seventy participants (32 with CLBP and 38 matched controls) completed a baseline MRI. Twelve of those with CLBP underwent an additional follow-up MRI, which is the focus of this study. Demographic information and Oswestry Disability Index scores for each participant are provided in Table 1.

**Table 1.** Demographic information about the study group.

| Participant ID | time to follow-up | age at baseline | sex | BMI | ODI baseline | ODI follow-up |
| --- | --- | --- | --- | --- | --- | --- |
| 1 | 36 | 46-50 | F | 25.9 | 48 | 34 |
| 2 | 31 | 40-45 | F | 24.4 | 30 | 58 |
| 3 | 27 | 20-25 | F | 28.2 | 48 | 10 |
| 4 | 24 | 30-35 | M | 26.4 | 32 | 22 |
| 5 | 24 | 40-45 | F | 23.0 | 48 | 32 |
| 6 | 23 | 30-35 | F | 27.4 | 30 | 20 |
| 7 | 27 | 40-45 | F | 28.0 | 55 | 52 |
| 8 | 20 | 36-40 | F | 32.4 | 22 | 22 |
| 9 | 14 | 56-60 | F | 38.7 | 58 | 13 |
| 10 | 14 | 50-55 | F | 27.9 | 22 | 16 |
| 11 | 32 | 50-55 | M | 28.1 | 0 | 0 |
| 12 | 10 | 60-65 | F | 22.9 | 33 | 16 |
| **Median** | 24 | 43 |  | 27.4 | 32.5 | 21 |
| **Mean ± SD** | 23.5±7.9 | 42±11.6 | | 27.2 | 35.5±16.7 | 24.6±16.9 |

Individuals with CLBP were referred to this study by healthcare providers within our institution. Participants were interviewed, and their medical charts were reviewed. Those with disc herniation, metabolic bone disease, any surgery or trauma at the lumbar level, osteoporosis, spondylolisthesis, or malignancy were excluded. To minimize the effects of daily activities on disc physiology, participants were asked to avoid strenuous physical activity and rested in a supine position for 30 minutes before the MRI scans. Additionally, the timing of the baseline and follow-up scans was randomized, resulting in five participants having their baseline MRI later than the follow-up, and seven having it earlier.

### Assessment of physical disability due to low back pain

Since there is no objective measure of pain, condition-specific health status measures are commonly used to assess a patient’s low back pain condition [22, 23]. In this study, Oswestry Disability Index (ODI) [22] was used, which is a widely used outcome measure based on a patient’s self-report on how low back pain affects their daily physical activities. Participants filled out the ODI questionnaire right before the baseline and the follow-up MRI exams.

### MRI acquisition

Images of the lumbar spine were acquired using a 3T GE Discovery MR750 MRI system (Waukesha, WI USA) equipped with a CTL-spine coil. T1 weighted (T2W) scans were acquired in the sagittal plane with a Fast Spin-Echo (FSE) pulse sequence with TR=780ms, TE=8.2ms, Echo Train Length (ETL) of 3, 1.2 mm in-plane resolution, 3mm slice thickness. T2 weighted (T2W) MRI were acquired using a FSE with TR=4500ms, TE=104ms, ETL=24 and the same resolution as the T_1_W scans. Diffusion MRI were acquired using a single-shot spin-echo echo planar imaging with b=600s/mm^2^, and a reference scan, TR=2100 ms, TE=75 ms, NEX=12, 2.4 mm in-plane resolution and 3mm slice thickness. *T1ρ* data were acquired with an FSE using a spin-lock amplitude of 400Hz, spin-lock durations (TSL) of 0ms, 20ms, 40ms, 60ms and TR=1400ms and TE=60ms.

Disc measurements described in the next section are done by manually outlining the regions of interest.

### Quantitative MRI metrics

#### Disc Height Index (DHI)

A recently introduced normalized disc height measurement method [13], compensates for body size variations, enabling inter-participant comparisons. This technique normalizes disc height using adjacent vertebral body dimensions. To calculate DHI (Eq.1), manual measurements of proximal (PV) and distal (DV) vertebral body height and disc height (DH) were taken from the anterior, middle, and posterior portions of each disc level on midsagittal slice on T2 weighted MRI images.

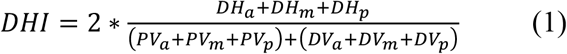

#### Apparent Diffusion Coefficient (ADC) calculation

ADC is a measure of bulk diffusion of water molecules, which is shown be a reliable measure of the degree of disc degeneration [14–18]. It is an indirect measure of pathologic changes in the nucleus pulposus (NP), such as dehydration, development of fibrocartilage and clusters of chondrocytes. The average ADC values were calculated inside the NP using Eq.2:

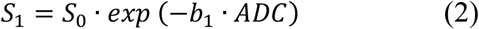

Here S_1_ and S_0_ are the signal intensities measured with and without the diffusion weighting, respectively.

#### T1ρ relaxation calculation

*T1ρ* (spin-lattice relaxation in the rotating frame) is a potential biomarker of changes in disc physiology. *T1ρ* values in the NP decrease with the loss of the proteoglycan (PG) content of the disc extracellular matrix [19, 24–27]. *T1ρ* values were calculated by fitting a mono-exponential model to the voxel signal with a series of spin-lock pulse durations (TSL):

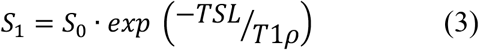

Here S_1_ and S_0_ are the signal intensities measured with and without the spin-locking RF pulse, respectively.

### Statistical Analysis

SPSS software version 26 and R Studio (Version 2023.12.0+369) were used for statistical analyses (Armonk, NY USA). A significance level of 0.05 (after adjusting for multiple comparisons using Bonferroni correction) was set for all analyses.

#### Correlation between MRI metrics and ODI

The aim was to examine whether similar changes occurred in MRI metrics from baseline to follow-up and if these changes were associated with ODI scores. For each participant, symmetrized percentage changes [28] were calculated between follow-up and baseline scans for each MRI metric and ODI score. Spearman’s rank correlation coefficient was then computed between each pair of these measures, performed for each lumbar disc separately.

#### Changes in MRI metrics between baseline and follow-up

A paired t-test was used to assess group-level changes in each MRI metric for each disc from baseline to follow-up, conducted separately for each lumbar disc.

### Comparing a disc with respect to normative values

Jarman et al. demonstrated that quantitative MRI measurements indicated accelerated degeneration when the DHI of a disc was more than 1.5 standard deviations below the average DHI of healthy discs (grades 1–2) [13]. Therefore, the discs of a participant were evaluated at baseline and follow-up by normalizing the DHI of each disc with respect to the mean and standard deviation of healthy discs at the respective level:

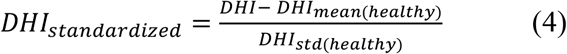

The means and standard deviations of DHI for the healthy discs are presented in Table 2, which were derived by selecting only discs with Pfirrmann grades 1 or 2 from the full set of 70 participants in the original study. Eq.4 is calculated for each disc using the respective values from table 2. The same calculation is repeated for average ADC and *T1ρ* in each disc.

**Table 2.** Mean and standard deviation of DHI for healthy discs (grades 1 – 2)

|  | L1/L2 | L2/L3 | L3/L4 | L4/L5 | L5/S1 |
| --- | --- | --- | --- | --- | --- |
| Mean DHI | 0.303 | 0.345 | 0.368 | 0.424 | 0.381 |
| Std. Dev. DHI | 0.048 | 0.051 | 0.061 | 0.062 | 0.061 |

The standardized scores provide a quantitative representation of whether a participant’s disc measurements fall within the healthy range. Additionally, these measurements serve as indicators of any measurable changes that may have occurred during a follow-up examination.

## RESULTS

### Correlation between changes in MRI metrics and changes in ODI

Results of the correlation analyses are shown in Table 3. There were strong positive correlations between DHI and ADC changes in L4/L5, ADC and *T1ρ* changes in L2/L3, and DHI and *T1ρ* changes in L5/S1. Only the latter was close to the statistical significance of p<0.0042 with the conservative Bonferroni correction.

**Table 3.** Results from the correlation analysis. This analysis explored if a change from baseline (BL) to follow-up (F/U) in one metric correlated with the others. Each cell lists the Spearman’s rank correlation coefficient, R (and p-values) for the respective analysis. Notable correlations before Bonferroni correction are marked with *. Strongest correlation was between disc height change and T1ρ change in L5/S1 disc.

| Disc | DHI<sub>BL-F/U</sub> vs<br>ADC<sub>BL-F/U</sub> | DHI<sub>BL-F/U</sub> vs<br>T1ρ<sub>BL-F/U</sub> | ADC<sub>BL-F/U</sub> vs<br>T1ρ<sub>BL-F/U</sub> | ODI<sub>BL-F/U</sub> vs<br>ADC<sub>BL-F/U</sub> | ODI<sub>BL-F/U</sub> vs<br>T1ρ<sub>BL-F/U</sub> | ODI<sub>BL-F/U</sub> vs<br>DHI<sub>BL-F/U</sub> |
| --- | --- | --- | --- | --- | --- | --- |
| L1/L2 | 0.14 (p=0.7) | -0.44 (p=0.15) | 0.27 (p=0.39) | -0.02 (p=0.94) | 0.21 (p=0.5) | 0.53 (p=0.08) |
| L2/L3 | -0.08 (p=0.8) | -0.18 (p=0.6) | 0.65* (p=0.02) | 0.47 (p=0.12) | 0.5 (p=0.1) | 0.68* (p=0.015) |
| L3/L4 | -0.47 (p=0.12) | -0.4 (p=0.2) | -0.45 (p=0.14) | 0.32 (p=0.3) | 0.32 (p=0.3) | -0.5 (p=0.1) |
| L4/L5 | 0.58* (p=0.05) | -0.15 (p=0.6) | 0.45 (p=0.14) | 0.39 (p=0.2) | 0.66* (p=0.02) | -0.32 (p=0.3) |
| L5/S1 | 0.25 (p=0.43) | **0.75\* (p=0.005)** | 0.18 (p=0.58) | 0.31 (p=0.3) | 0.36 (p=0.25) | 0.12 (p=0.7) |

### Changes in MRI metrics between baseline and follow-up

Paired t-test results are listed in table 4. All lumbar discs had significant disc height loss between the baseline and follow-up scans. If stringent Bonferroni correction was used to account for multiple comparisons, changes in L2/L3 and L4/L5 were not significant at p<0.01. There were no measurable changes in *T1ρ* or ADC. Also, the changes in ODI varied from subject to subject (p=0.066). Fig.1 shows the boxplots for the three MRI metrics for each lumbar disc at baseline and follow-up.

**Fig 1.**
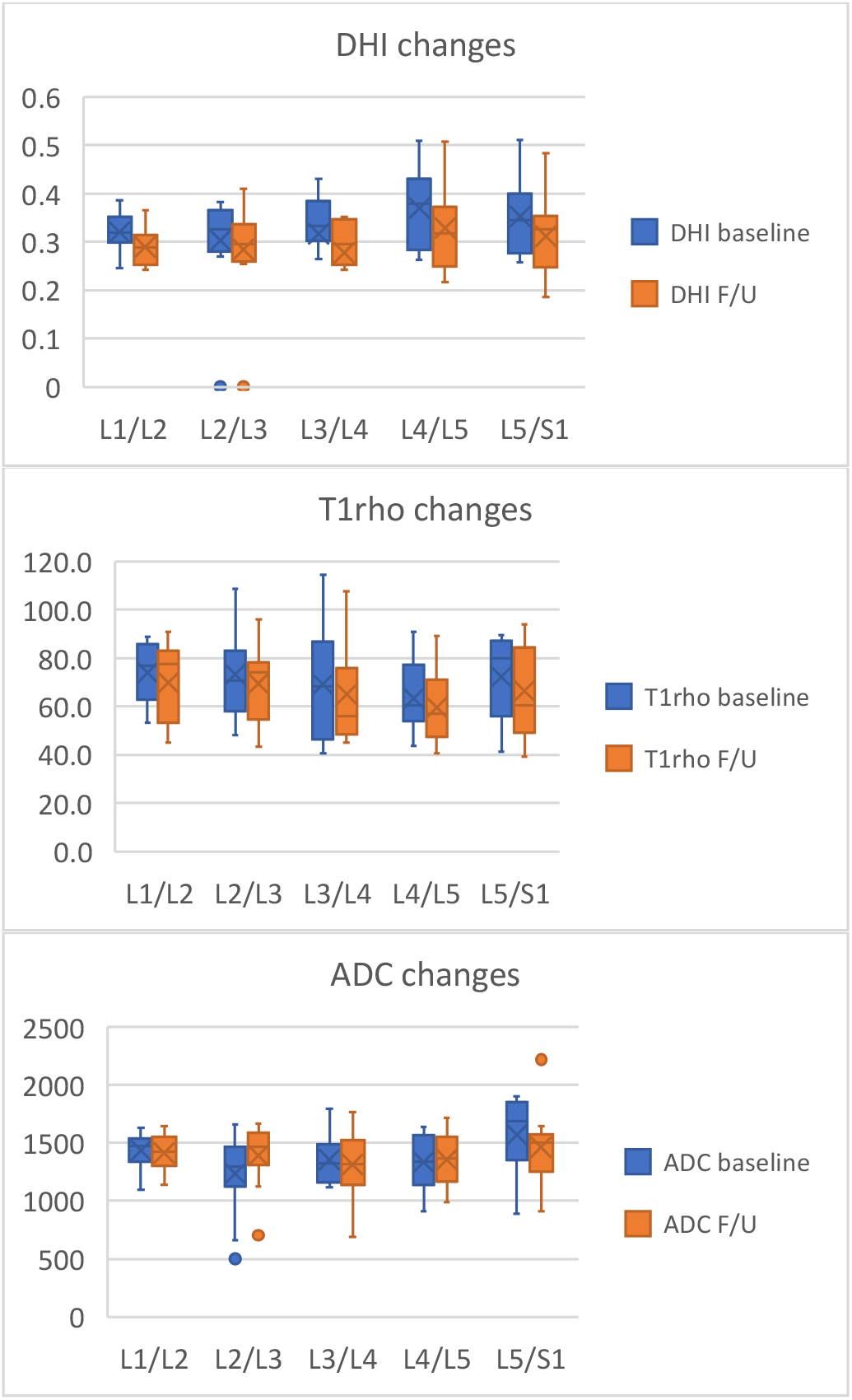
Boxplots for DHI, T1ρ and ADC at baseline and follow-up scans for each lumbar disc. There were significant losses in disc height at all levels, while other metrics did not show such clear trends.

**Table 4.** Results from the paired t-test analyses that explored changes in quantitative MRI metrics in each lumbar disc. T-scores (and p-values) are reported in each cell. Statistically significant results after Bonferroni adjustment are marked with bold letters and *.

|  | DHI change | *T1ρ* change | ADC change |
| --- | --- | --- | --- |
| L1/L2 | **4.43\* (p=0.001)** | 1.6 (p=0.14) | 0.43 (p=0.67) |
| L2/L3 | 2.3 (p=0.044) | 1.7 (p=0.11) | -2.8 (p=0.016) |
| L3/L4 | **4.35\* (p=0.0012)** | 1.6 (p=0.13) | 0.64 (p=0.53) |
| L4/L5 | 3.13 (p=0.009) | 1.06 (p=0.31) | -0.59 (p=0.56) |
| L5/S1 | **4.14\* (p=0.0016)** | 1.3 (p=0.23) | 1.16 (p=0.27) |

#### Inter-subject variations in DHI loss

The extent of DHI loss differed among participants. Fig. 2 illustrates the distribution of percent DHI changes between baseline and follow-up across all discs from all participants. As shown in the figure, DHI changes in this cohort were generally around 15% to 20%, with some reaching up to 50% (median is 10% and standard deviation is 11%).

**Fig 2.**
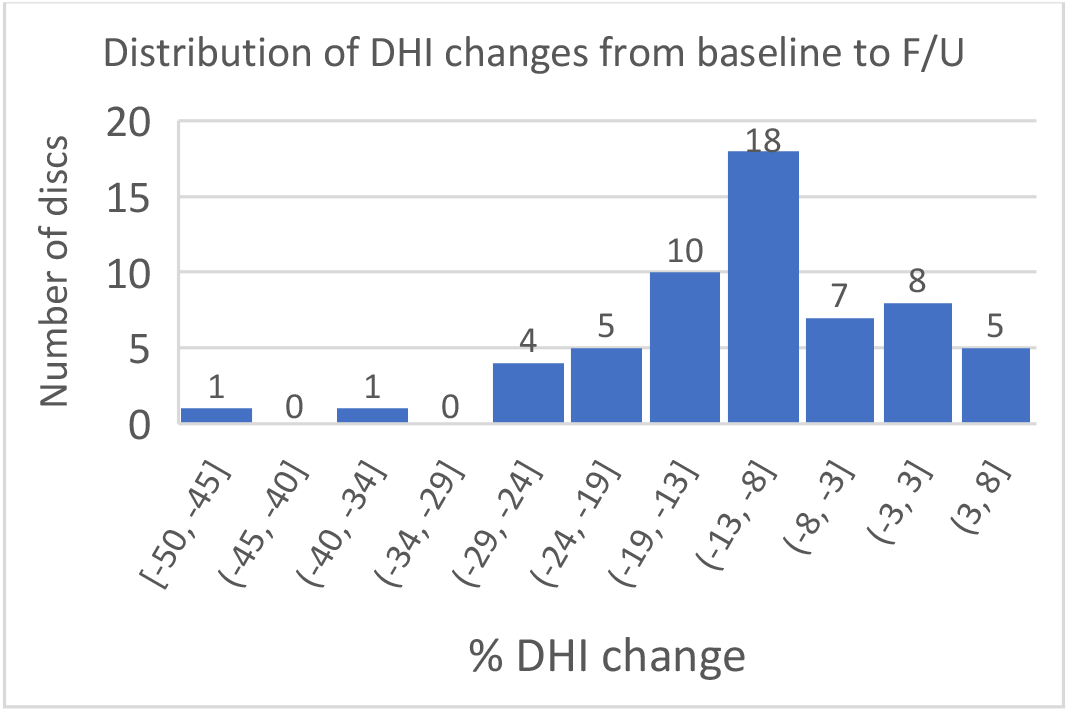
Histogram of percentage DHI changes from the baseline to the follow-up for all 60 lumbar discs from the 12 participants. The lower and upper limits of each bin is listed in the horizontal axis. For example, 16 discs had DHI loss between −11.49% and −5.03%.

#### Participant specific changes in quantitative MRI metrics

Data from one of the participants is provided to demonstrate the use of quantitative MRI metrics to assess individual patients. The top panel in Fig.3. shows the mid-sagittal slices from the baseline and follow-up T2W MRIs for one participant. The three plots below the MRIs show standardized DHI scores, *T1ρ* and ADC in units of standard deviation for each lumbar disc. A negative value of −1.5 or more indicates that a disc has values outside of healthy discs at that level [13]. These plots are useful for determining if a disc had healthy values at baseline and if it showed signs of degeneration between baseline and follow-up (adjacent blue and orange bars). Although visual inspection suggests a minor decrease in disc height only in L4/L5 disc, DHI metric showed that both L4/L5 and L5/S1 had measurable decrease in disc height. Moreover, all discs showed 25% - 42% decrease in *T1ρ* values.

**Fig 3.**
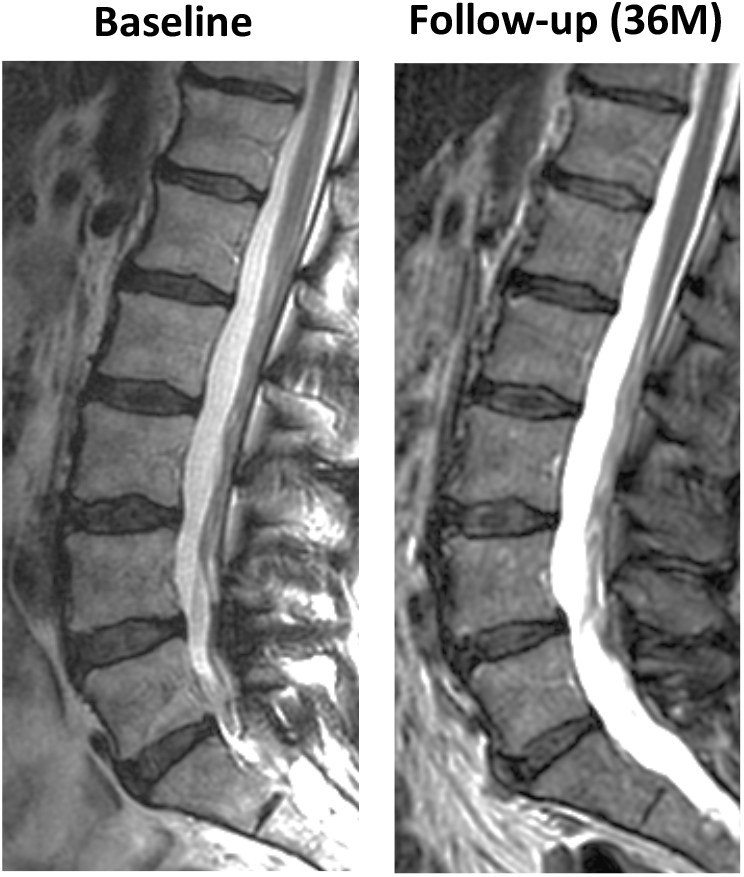
T2W scans from one of the participants and corresponding MRI metrics at baseline and follow-up. DHI, average T1ρ and ADC values are normalized with the mean and standard deviation obtained from the healthy discs (grades 1 and 2). Blue and orange bars show respective measurements at baseline and follow-up. DHI plots show that all discs were below the mean, but L1/L2 through L3/L4 remained relatively stable. On the other hand, L5/S1 underwent the largest amount of height loss. The time between the two scans was 36 months for this participant

**Fig 4.**
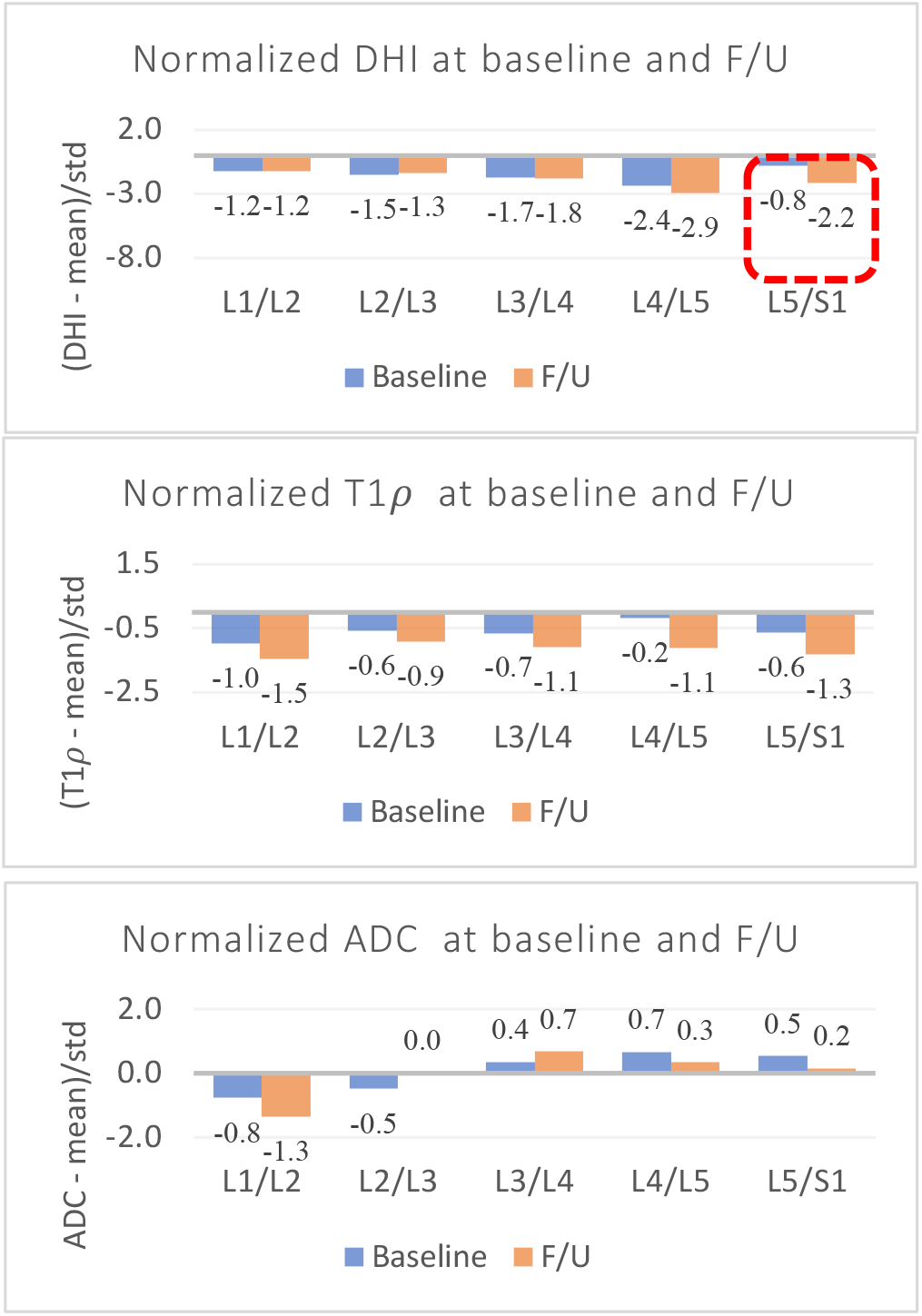
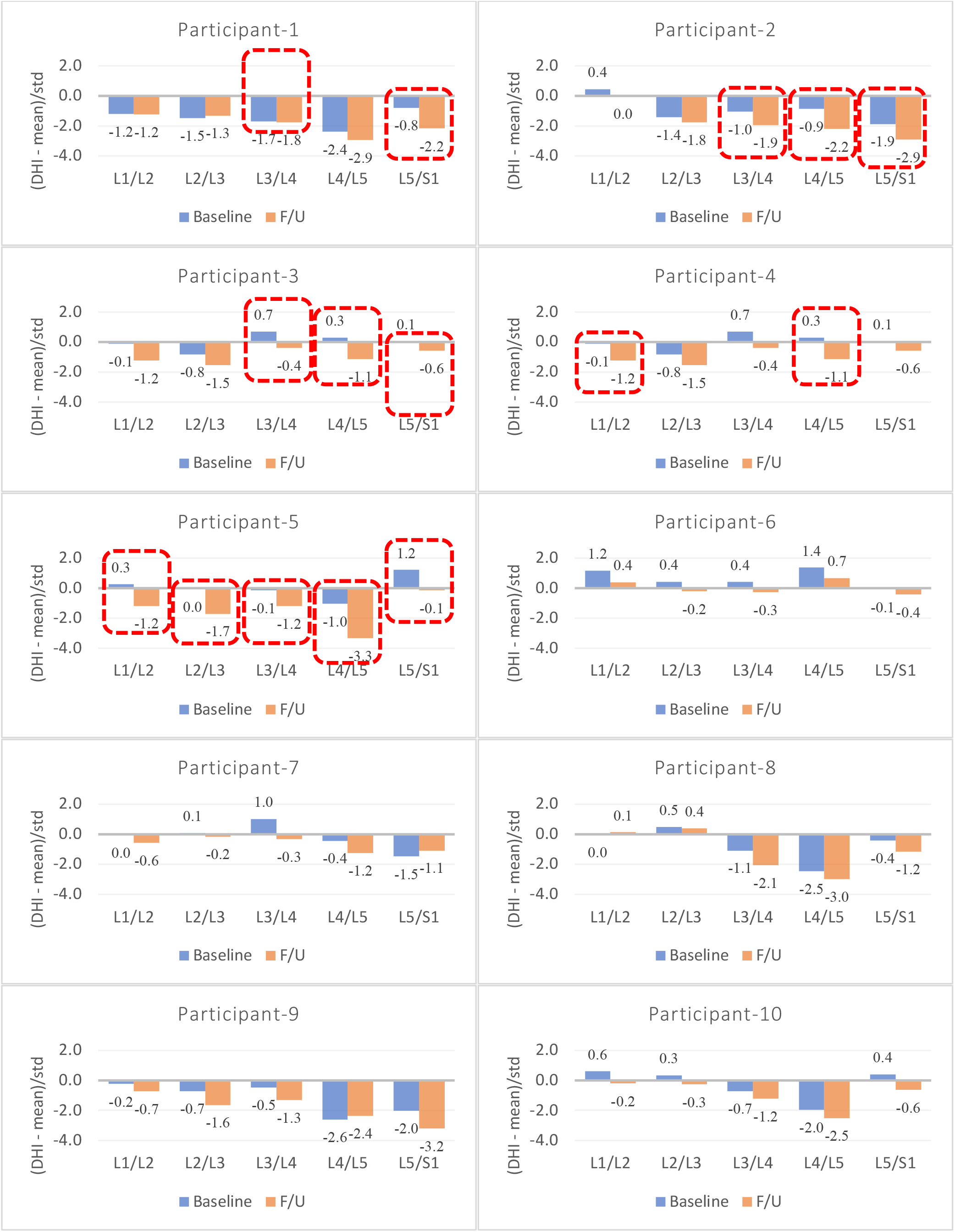
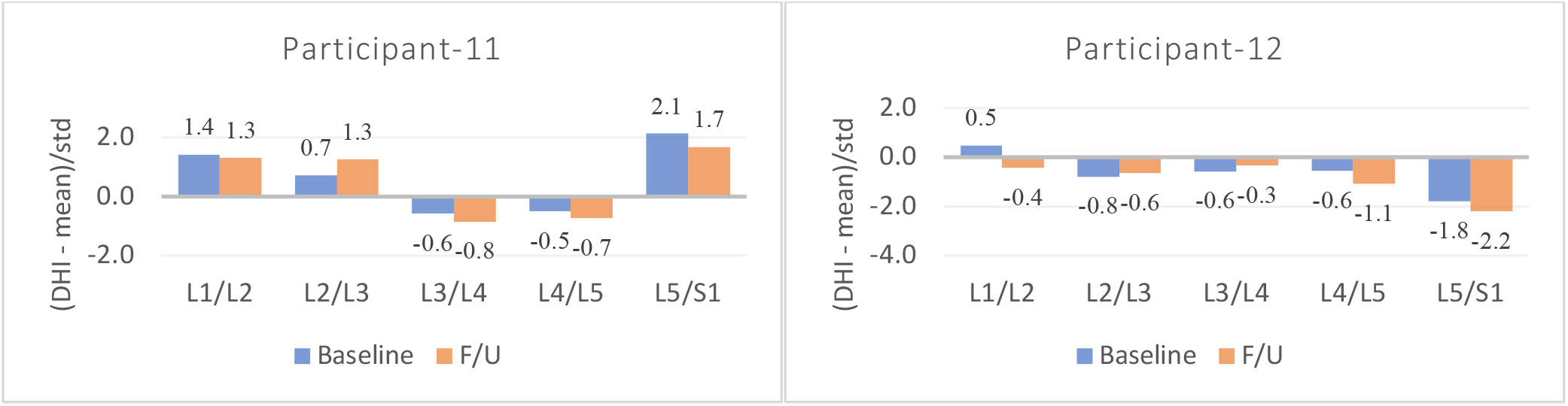
DHI (expressed in standard score, Eq. 4) of five lumbar discs at baseline and follow-up for each participant. The height of bars represents the DHI of the disc with respect to the healthy discs (in units of standard deviation in healthy discs). The discs that lost one standard deviation or higher is enclosed in red boxes. The mean and standard deviation of DHI used for standard score is calculated from all healthy discs (grades 1-2) from the full cohort of 70 participants.

## DISCUSSION

This study explored longitudinal changes in three quantitative MRI metrics and ODI in twelve participants. Some of the participants showed significant loss in DHI during a period of approximately two years, while others remained stable. This could indicate pathologic changes in respective discs beyond the typical age-related degeneration.

One of the strengths of this study was the ability to show changes in individual patients quantitatively, compared to earlier studies that focused on group-level changes using qualitative assessments. Group-level statistics were provided to show typical changes from baseline to follow-up in a relatively short time interval. In addition, the results from one of the participants were shown to illustrate the potential of such quantitative techniques to aid in clinical assessments. Although the two MRI scans looked similar qualitatively for the participant shown in Fig.3, complementary information in the plots show measurable changes. First, this participant had lower DHI and *T1ρ* even at baseline compared to healthy discs. Moreover, L5/S1 showed rapid height loss from 0.8 to 2.2 standard deviations below the mean of healthy discs. Low *T1ρ* values across all discs suggest that the discs might already be undergoing some physiological changes (loss of proteoglycans) and some further progression was noted.

The distribution of DHI loss in the 60 discs from the 12 participants show the disc height variations in this cohort (Fig.2). Half of the discs had a DHI loss between 5% - 18%, while some lost as much as 50%. Nine discs had slight positive change, which could be attributed to measurement errors.

While most participants experienced various degrees of disc height loss, other MRI metrics did not show such clear trends in all discs. L5/S1 showed the strongest correlation between disc height loss and decreasing *T1ρ*. This suggests that the structural and physiological changes do not always take place simultaneously during different stages of degeneration.

### Baseline Follow-up (36M)s

Our study was designed to minimize the effects of daily physical activities on disc physiology. Moreover, the changes that we observed between baseline and follow-up were much larger than those reported in a recent study of diurnal changes in disc height, volume and *T1ρ* conducted on 12 participants [30]. Their first scan was consistently at 7:00 in the morning (with 45 min rest before scans, which is close to ours) and the second one was done at 15:30 with no rest. They reported 6% - 8% height changes. Note that their participants did not rest for the afternoon scans, while our participants rested on the stretcher before both MRIs to reduce diurnal changes.

Although the size of the cohort was small, our approach sets up a foundation on which we can keep building as we acquire more data. Note that supplementary data are provided to show DHI at baseline and follow-up for all participants. Note that a similar study on diurnal changes in discs also studied only twelve participants and reported robust results [30]. The findings here suggest intriguing evidence of measurable changes in MRIs in about 2 years.

Another limitation was the gender imbalance. Although disc size normalization in DHI calculation (Eq.1) compensated for body size differences, other sex-related differences were not considered.

While we used hand-drawn measurements performed earlier in the study, we are currently working on automating the workflow using Machine Learning tools. This will allow physicians to receive the information immediately after scans are completed, thus streamlining the clinical process.

## Data Availability

All data produced in the present study are available upon reasonable request to the authors

